# Seroprevalence and Determinants of SARS-Cov-2 Antibodies Among Healthcare Workers in Dar es Salaam, Tanzania

**DOI:** 10.1101/2025.02.12.25322138

**Authors:** Erick Mboya, Claudia Hawkins, Doreen Kamori, Davis Amani, Grace Shayo, Raphael Z. Sangeda, Joan Rugemalila, Faraja Chiwanga, Sabina Mugusi, Upendo Nkwera, Mangaro Mabusi, Said Aboud, Ferdinand Mugusi, Bruno Sunguya, Muhammad Bakari, Tumaini Nagu

## Abstract

**Background:** Studies from the Sub-Saharan African (SSA) countries suggest an underestimate of SARS-CoV-2 infections early in the pandemic. To understand and inform infection control strategies to protect key and vulnerable populations, this study investigated the seroprevalence and factors associated with anti-SARS CoV-2 seropositivity among healthcare workers (HCWs) in Dar es Salaam, Tanzania.

**Methods:** This cross-sectional study was conducted in three tertiary and national hospitals in Dar es Salaam, Tanzania. Employed HCWs aged ≥18 years were enrolled between October 2021 and March 2022. Laboratory testing for Immunoglobulin G (IgG) against SARS-CoV-2 nucleocapsid (N) or spike (S) proteins was performed using WHO/FDA pre-qualified SARS-CoV-2 ELISA test kits, and data were collected on demographics and COVID-19-related exposures. Descriptive statistics were used to determine the SARS-CoV-2 antibody prevalence. Multivariable Modified Poisson regression with robust standard errors accounting for clustering within sites was used to test the independent association between HCW characteristics and SARS-CoV-2 serologic status.

**Results:** A total of 402 participants [mean age (SD) of 37.9 (9.7) years] were included in this study. 150 (37.3%) reported receiving at least one dose of SARS-CoV-2 vaccine. The overall seroprevalence of anti-SARS-CoV-2 antibodies (anti-SARS-CoV-2 IgG (S or N)) was 90.9% (358/394) among those with valid results. HCWs who reported receiving the SARS-CoV-2 vaccine were 1.14 times more likely to have anti-SARS-CoV-2 antibodies compared to those who reported not being vaccinated (aPR (95% CI) = 1.14 (1.03- 1.26)). HCWs who believed to have contracted COVID-19 previously but never tested had an 11% higher risk of being seropositive for SARS-CoV-2 IgG compared to those who thought never to have contracted COVID-19 before (aPR (95% CI) = 1.11 (1.05- 1.16)). Anesthetists and HCWs working in the laboratory and Obstetrics and Gynecology departments were more likely to have SARS-CoV-2 antibodies than HCWs from the Emergency Medicine Department (EMD).

**Conclusion:** The seroprevalence of anti-SARS-CoV-2 IgG among HCWs in this study was high in this population indicating high SARS-CoV-2 exposure among HCWs. Our work highlights the need for more effective infection control practices in healthcare settings in future pandemics like these, especially among HCWs at highest risk.

## Background

The COVID-19 pandemic placed a significant strain on healthcare systems worldwide (1,2). Low- and middle-income countries (LMICs) faced distinct challenges during the COVID-19 pandemic such as limited healthcare, testing, and vaccine access and acceptability (3–5). Healthcare workers (HCWs) on the frontline in the fight against SARS-CoV-2, were also impacted profoundly during the pandemic. Increased occupational exposure to viral particles, inadequate protective equipment and infection control practices, and other stressors such as long working hours, put them at a significantly higher risk of acquiring SARS-CoV-2 compared to the general population (6,7)(7). HCWs also reported experiencing higher workloads, psychological distress, social exclusion (2).

In the US and other settings, up to about 30% of the healthcare workforce was infected with SARS-CoV-2 during the peak of COVID-19 (8,9). According to the World Health Organization (WHO), approximately 115,500 HCWs died in 2021 due to COVID-19-related complications globally (10). Studies examining the prevalence of SARS-CoV-2 infection in LMICs are more limited due to a lack of accessible testing and surveillance (9). The limited data other Sub-Saharan African (SSA) countries suggests the burden of SARS-CoV-2 infections among HCWs was significantly underestimated. Like its neighboring countries, Tanzania imposed measures to prevent the spread of COVID-19 including face masks in densely populated areas, prohibiting large public gatherings, encouraging the use of sanitizer and installing hand-washing facilities in public places and households, assigning isolation and quarantine centers and expanded diagnostic points of service (11). However, Tanzania did not impose curfews, nor complete country lock-down, and encouraged income-generating activities (11,12). The lack of data on the burden and risks associated with infection significantly hinders understanding of the impact of SARS-CoV-2, especially in high-risk individuals like HCWs, and the ability to formulate evidence-based effective interventions for subsequent outbreaks (13).

This study investigated the seroprevalence and factors associated with anti-SARS CoV-2 seropositivity among HCWs in Dar es Salaam, Tanzania. This data is important, particularly in Tanzania to inform infection control and mitigation policies that best protect these vulnerable workers in future outbreaks.

## Methodology

### Study design and duration

This cross-sectional analysis was conducted among HCWs enrolled in a longitudinal study examining the seroprevalence and seroincidence of SARS-CoV-2 in three tertiary hospitals in Dar es Salaam; the Mloganzila Campus of the National Hospital (Mloganzila Hospital), the Muhimbili National Hospital (MNH), and Amana Regional Referral Hospital (ARRH). Recruitment occurred between October 2021 and March 2022. Specifically, at MNH recruitment was between October 6^th^ to November 22^nd,^ 2021, while at Mloganzila Hospital recruitment was between January 21^st^ to February 11^th,^ 2022, and February 22^nd^ to March 11^th,^ 2022 at Amana Regional Referral Hospital. Recruitment of participants Mloganzila and ARRH occured during the peak of the second wave of COVID-19 in the country including Dar es Salaam (Figure 1).

**Figure 1:**
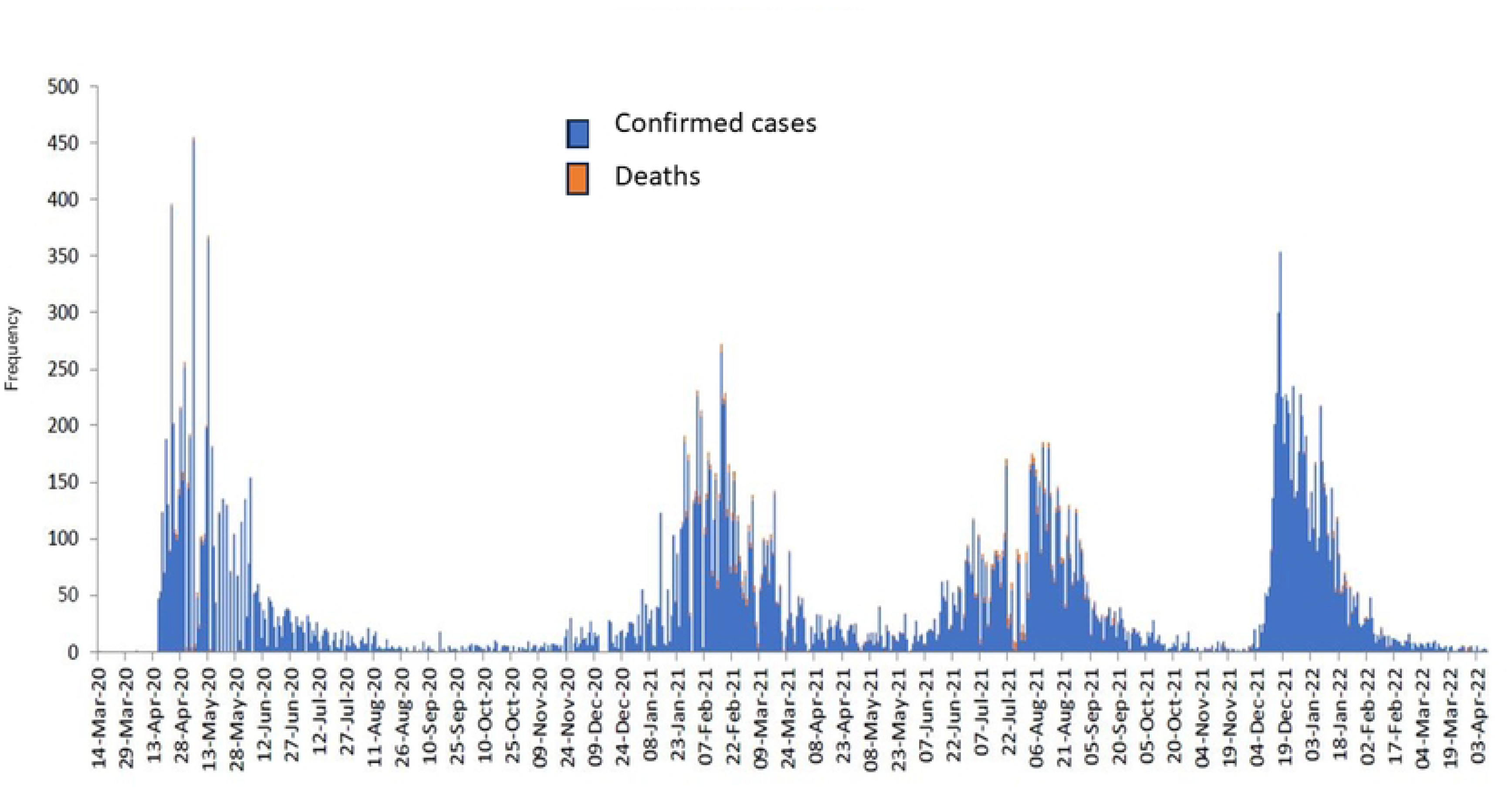
Trend of cases and deaths from COVID-19 since March 2020 to the week ending on 8^th^ April 2022. Source: Ministry of Health Tanzania COVID-19 Situation report Number 15 as of April 2022 (14)

### Study Setting

The first case of COVID-19 in Tanzania was imported on March 16^th^, 2020 (15). At the time of this study, there had been four waves of COVID-19 in Tanzania (Figure 1). As of February 2023, 40,000 cases of COVID-19 have been confirmed, mostly in Dar es Salaam (14). Johnson & Johnson vaccine was the widely available vaccine through the COVAX program, free of charge and HCWs were among the prioritized groups (16). Face masks in densely populated areas were recommended, although large public gatherings were not prohibited. Sanitizers and hand-washing facilities in public places were installed and there were assigned isolation and quarantine centers (11). The three hospitals included in this study are public hospitals and were designated COVID-19 centers where patients with severe COVID-19 from lower facilities in Dar es Salaam were referred for treatment and isolation.

### Study population

All HCWs 18 years and older working were eligible for inclusion in this study. Students, volunteers/interns, and temporary workers with contracts of less than 7 months left on their contract were excluded from the cohort.

### Sampling

Stratified random sampling was used to select participants. Participants were stratified by their institution, cadre, and department. Probability proportional to size was used to allocate the number for each stratum.

Since the estimate for the SARS-CoV-2 exposure was not known, 50% exposure was set to maximize the sample size for a finite population of 3,000 HCWs. Accounting for the non-response of 15% the sample size for this study was calculated to be 393 HCWs to estimate SARS-CoV-2 exposure with 5% absolute precision.

### Recruitment

Data on the number of employees were obtained from each institution and were stratified by their cadre (doctors, nurses, laboratory scientists, pharmacists, and allied health workers). Random selection of HCWs was performed using a lottery paper method and those selected were invited to participate in the study. Participants who provided a signed consent to the study were then enrolled and assigned unique study numbers. Participants had their weight and height measured and were administered questionnaires asking about demographics, occupational risks, reported COVID-19 infection, results of any testing, other COVID-19-related exposures, COVID-19 symptom severity, and COVID-19 vaccination details if received. Participants were then asked to provide a single blood sample of 8 ml volume for SARS-CoV-2 IgG testing, which was collected by a trained study staff.

### Laboratory procedures

Samples were tested by conventional standard ELISA platform at the MNH Central Pathology Laboratory (CPL), an ISO-certified laboratory, using WHO/FDA pre-qualified SARS-CoV-2 ELISA test kits, testing for SARS-CoV-2 IgG (nucleocapsid and spike). The results of the test were interpreted according to the manufacturer’s instructions (17).

The interpretation of the serological results was as follows (17):

a. Positive for both anti-N and anti-S antibodies

i. COVID-19 vaccinated +/- previous SARS-CoV-2 infection.
ii. unvaccinated with previous SARS-CoV-2 infection.
b. Positive anti-N antibody and negative anti-S antibody

i. unvaccinated with previous SARS-CoV-2 infection.
ii. COVID-19 vaccinated with previous SARS-CoV-2 infection but no response to the vaccine or loss of vaccine response.
c. Negative anti-N antibody and positive anti-S antibody

i. COVID-vaccination with no prior SARS-CoV-2 infection.
ii. previous or current exposure to SARS-CoV-2 and/or COVID-19 vaccination
d. Negative anti-N antibody and negative anti-S antibody

i. No prior or current exposure to SARS-CoV-2 or COVID vaccination
ii. Previous or current exposure to SARS-CoV-2 and/or COVID vaccination with no immune response

### Data management and analysis

All data was entered electronically using REDCap version 11.1.5 (Vanderbilt University). Data were extracted in Stata format, cleaned, and analyzed using Stata 18. Participants’ characteristics were summarized using frequencies and percentages for all patients and disaggregated for each facility. Age was summarized using mean and standard deviation (SD). The overall seroprevalence of SARS CoV-2 (SARS-CoV-2 IgG) was calculated as the proportion of HCWs who tested positive for *either* IgG S or N among all HCWs with the serology results. The proportion of HCWs who tested positive for *both* IgG S and N or IgG S alone was also calculated and disaggregated by sites, vaccination status, and other demographic characteristics. Multivariable Modified Poisson regression with robust variance estimators accounting for clustering within sites was used to test the independent association between HCWs characteristics and overall seroprevalence of SARS CoV-2 IgG among HCWs in Dar es Salaam. A significance level of <0.05 was used to determine a significant association. All factors that had a p-value of <0.2 in the univariable analysis were included in the multivariable model but were only kept in the final model if they improved the model. The area under the curve (AUC) was used to select a final model (AUC=0.8162).

## Results

### Characteristics of the study participants

A total of 402 HCWs were included in this study; [mean age (±SD) was 37.9 (±9.7) years; 236 (58.7%) females]. A quarter of the participants, 105 (26.1%) worked in the Internal Medicine department, 78 (19.4%) in the Obstetrics and Gynecology department, and 58 (14.4%) from other surgical departments. By their cadres, 174 (42.5%) were nursing staff, followed by medical officers/dentists 80 (19.6%) and specialists 61 (14.9%). About a fifth of the HCWs, 71 (17.7%), reported spending more than half of their time working in the ICU (**Table 1**).

**Table 1:**
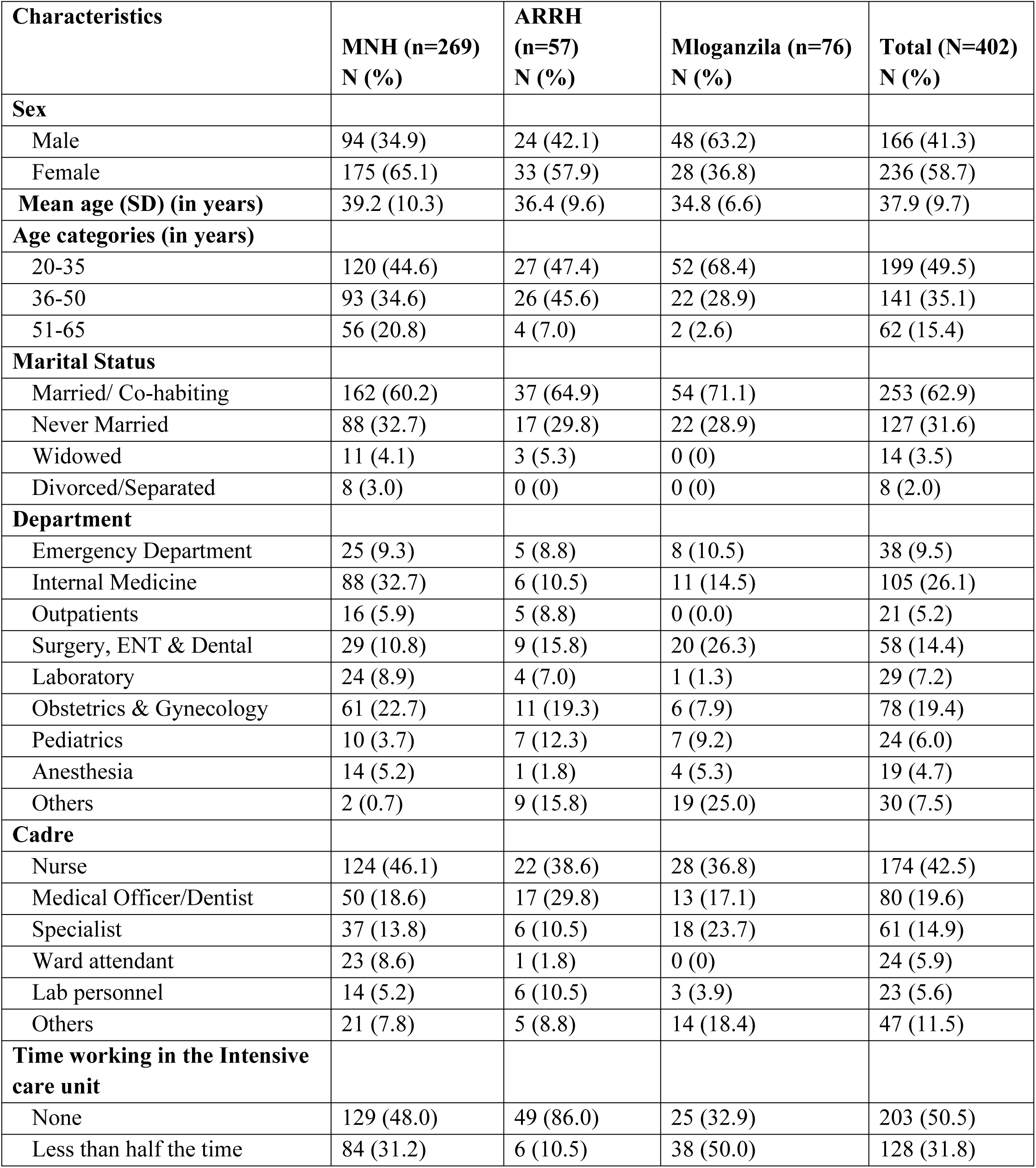
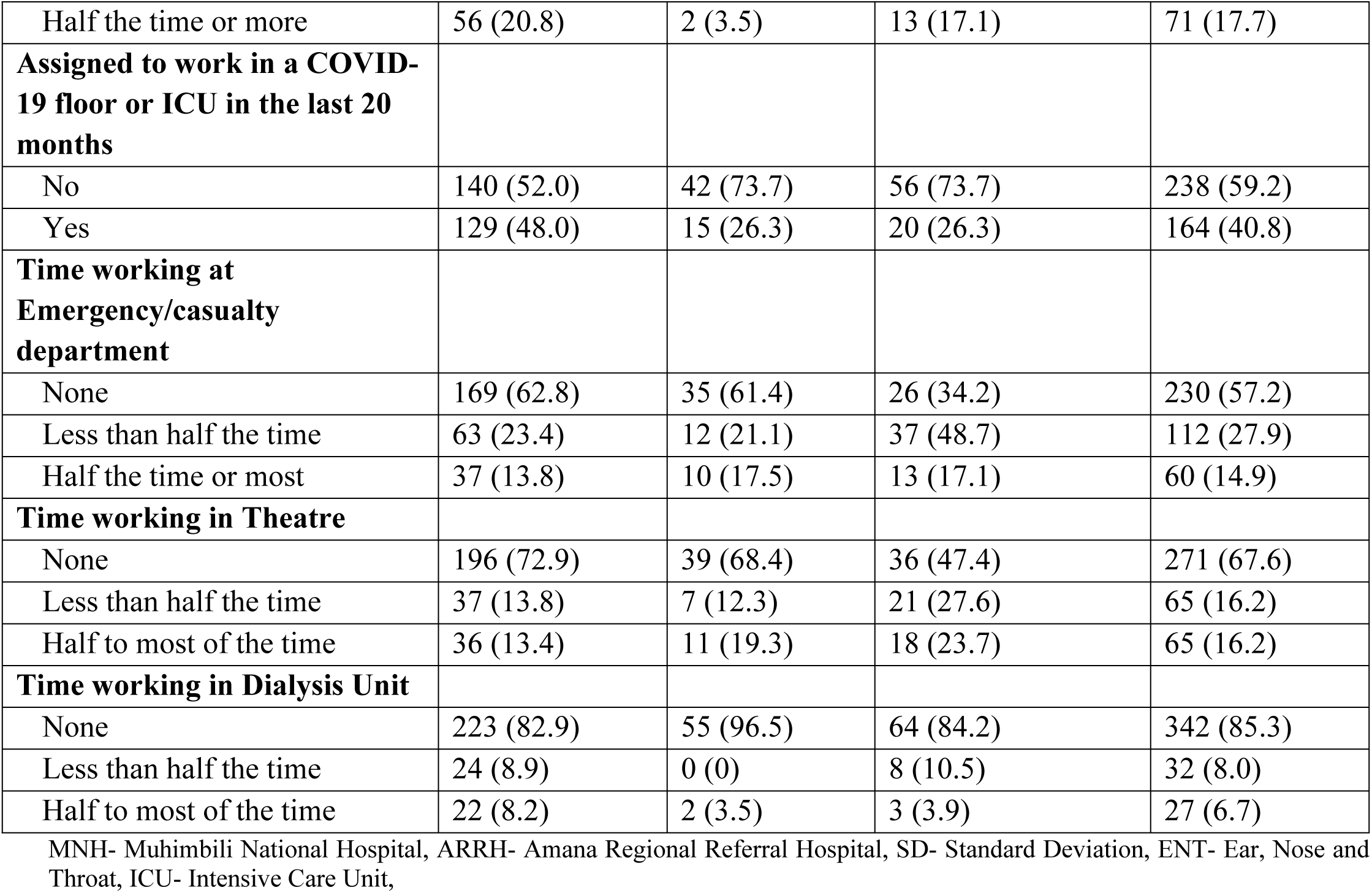
Demographic and occupational characteristics of HCWs by site (N=402)

Sixty-nine participants (17.2%) reported previous testing for COVID-19 and among them 21 (30.4%) tested positive. Twenty-one (5.2%) participants reported to have previously been admitted to the hospital for COVID-19; with 6 of them (28.6%) admitted for less than a week. Among all participants, 150 (37.3%) reported to have received at least one dose of a vaccine against COVID-19; 94% received a dose of the Johnson & Johnson vaccine, **Table 2**.

**Table 2:**
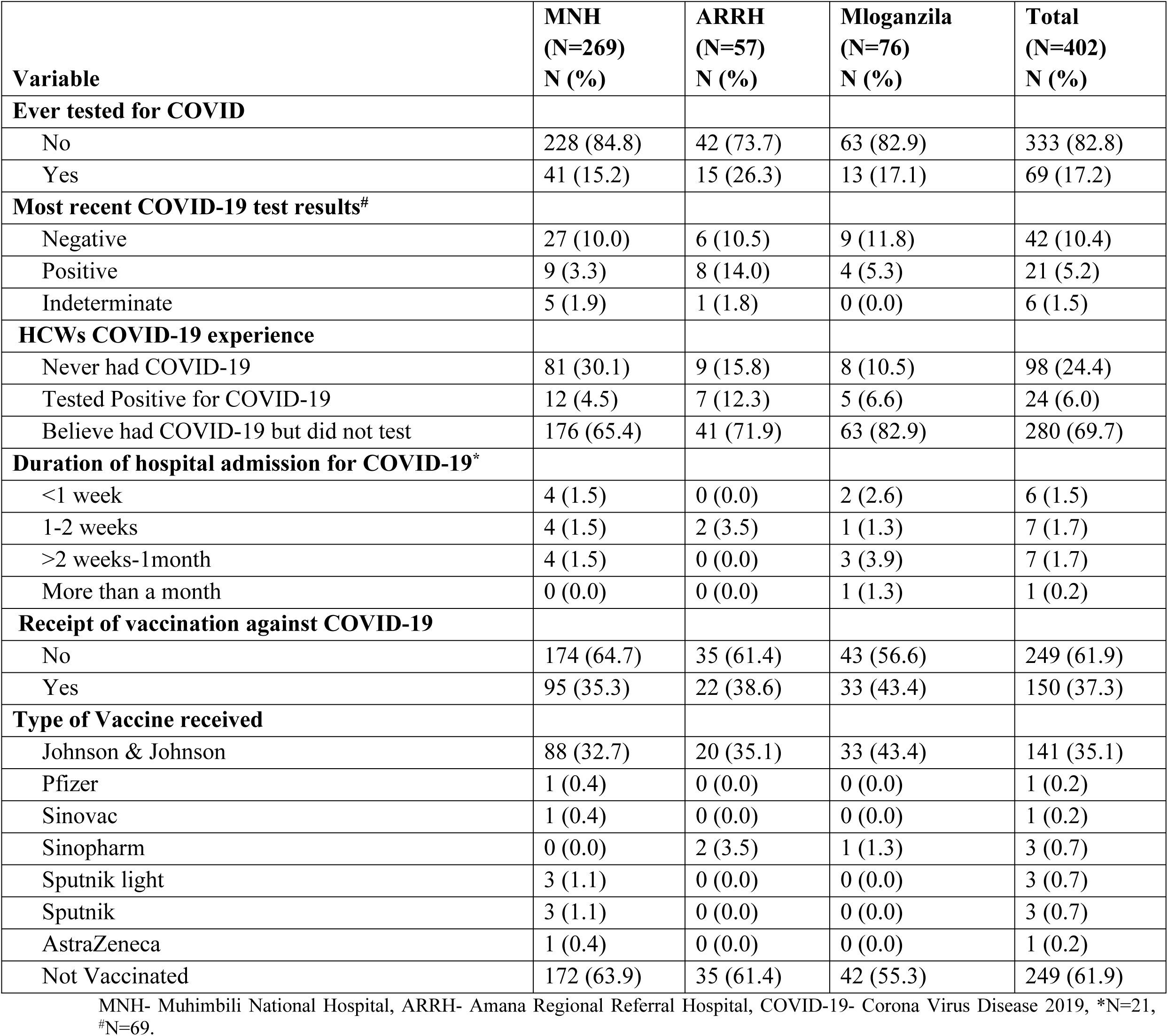
COVID-19 experience, testing, and vaccination among HCWs in Dar es Salaam (N=402)

### Seroprevalence of total SARS-CoV-2 Ig-G among HCWs in Dar es Salaam

Among the 402 HCWs, serology results were available for analysis for 394 HCWs. Of the 8 participants excluded, 6 had missing results and 2 did not have a unique identifier to link the results with the electronic clinical data.

The overall seroprevalence of SARS-CoV-2 IgG (IgG N, IgG S, or both IgG N and S) among HCWs was 91.8%. The overall seroprevalence by site was 89.4%, 98.1%, and 95.8% among HCWs at MNH, Mloganzila and ARRH respectively (Figure 2).

**Figure 2:**
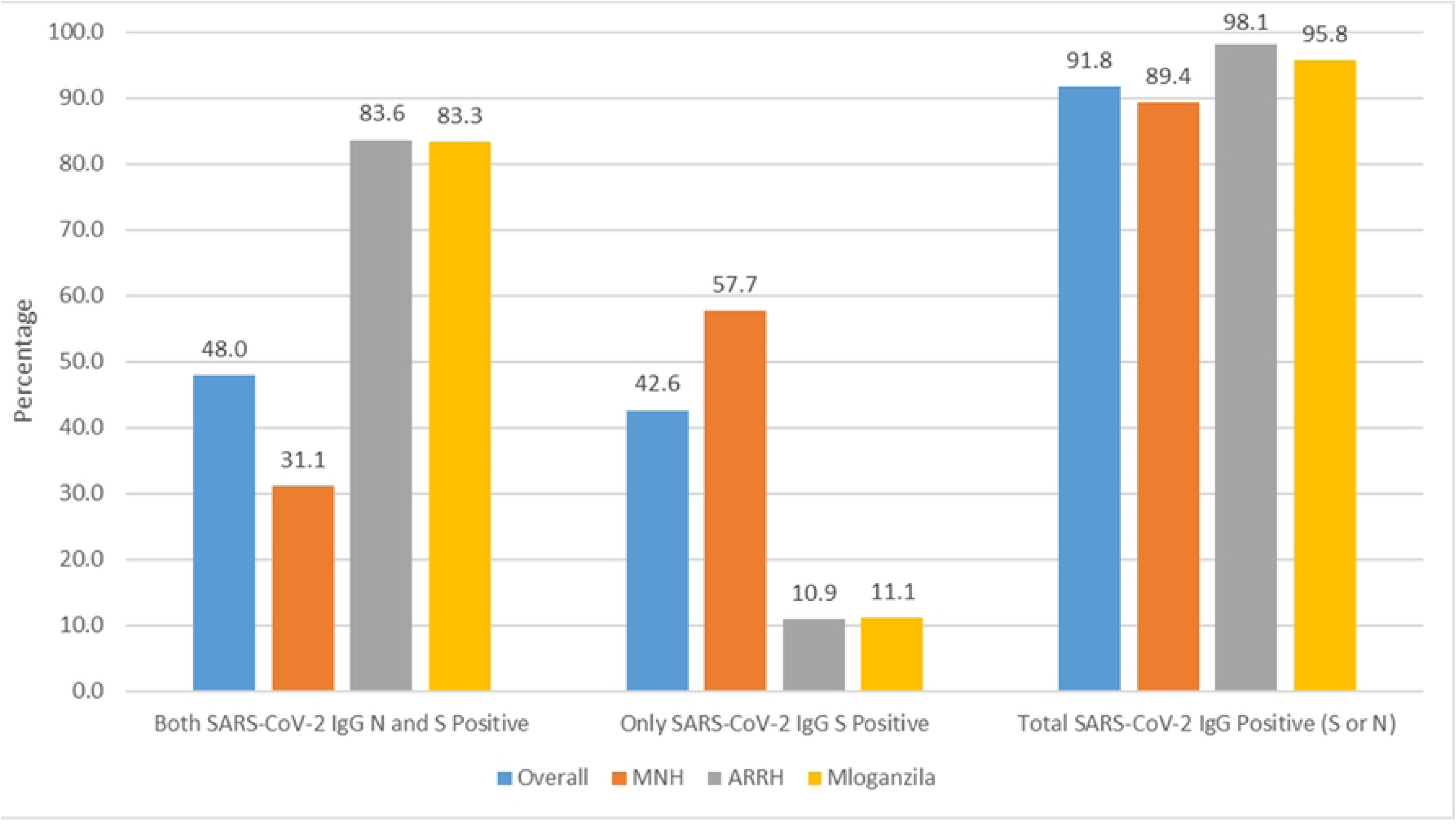
Prevalence of SARS-CoV-2 IgG by Site.

Almost half of the HCWs, 189 (48.0%), tested positive for *both* S and N SARS-CoV-2 IgG antibodies. SARS-CoV-2 IgG antibodies against both the N and S proteins were detected among 46 (83.6%) and 65 (83.3%) HCWs at Mloganzila hospital and ARRH respectively. At MNH, only (83 (31.1%)) HCWs tested positive for both the S and N SARS-CoV-2 IgG (**Figure 2)**.

Overall, 168 (42.6%) HCWs tested positive for SARS-CoV-2 IgG against the S protein only. At the MNH, 151 (57.7%) were positive for SARS-CoV-2 IgG S alone, while 6 (10.9%) HCWs at ARRH and 8 (11.1%) at Mloganzila hospital were positive against S protein alone. **(Figure 2).**

Among HCWs who reported to have received at least one dose of vaccine against COVID-19, the total seroprevalence of SARS-CoV-2 IgG was 97.8%. Among HCWs who reported not to have received the COVID-19 vaccine the seroprevalence of SARS-CoV-2 IgG was 86.6%. Similar rates of both SARS-CoV-2 IgG S and N positivity were present among vaccinated, 46.0%, and unvaccinated HCWs, 47.0% (Figure 3).

**Figure 3:**
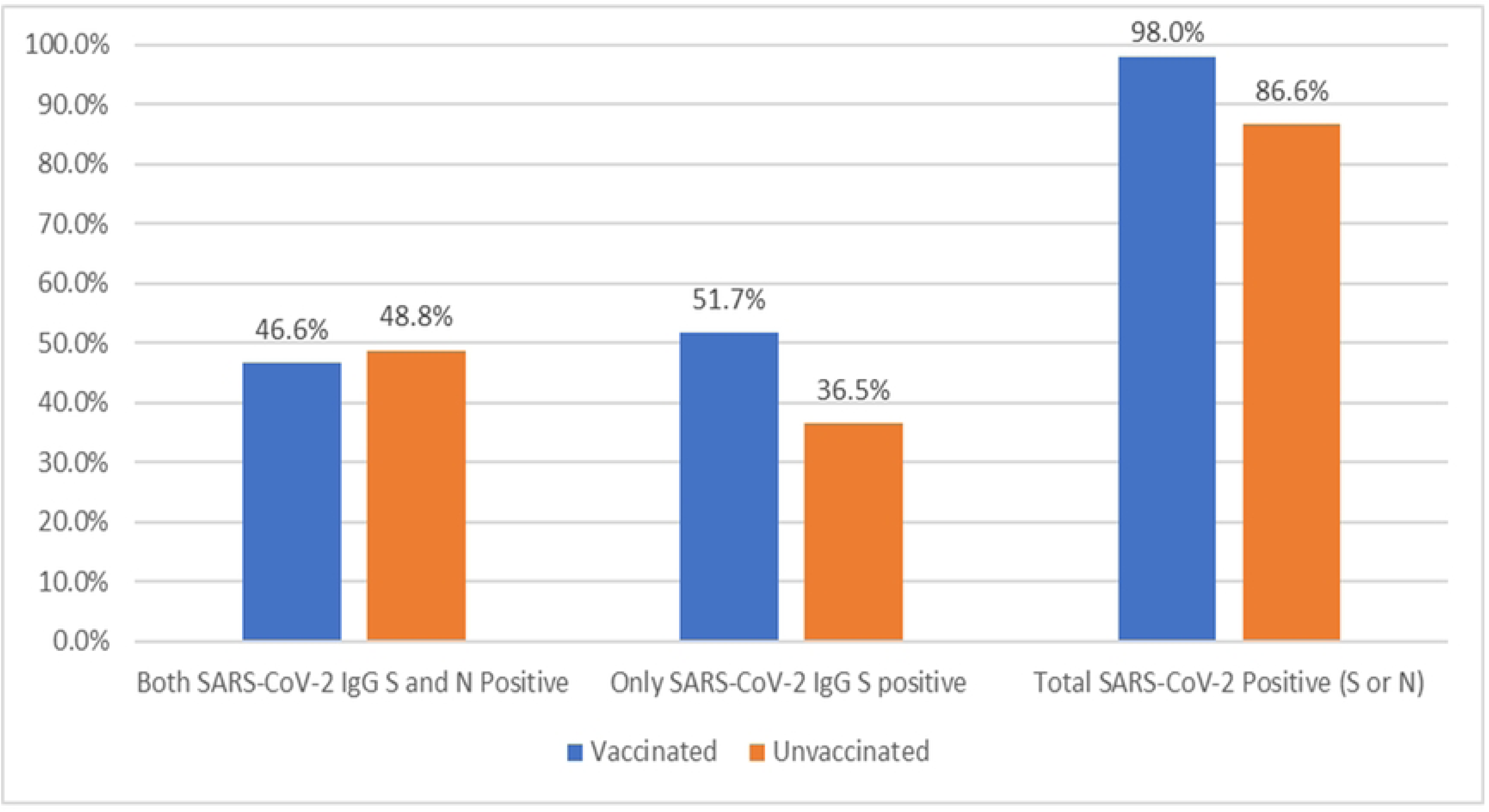
Prevalence of SARS-CoV2 IgG by COVID-19 Vaccination status.

Among vaccinated HCWs, 50.7% had IgG S only while only 36.5% of the unvaccinated had IgG S only (Figure 3).

### Factors associated with SARS-CoV-2 IgG seropositivity (IgG S or N positivity)

After adjusting for sex, marital status, department, cadre, vaccination status, previous COVID-19, and site (hospital) the following factors were independently associated with the risk of being seropositive for SARS-CoV-2 IgG among HCWs in Dar es Salaam: vaccination against COVID-19, department, cadre, previous COVID-19, site (hospital), sex and marital status.

HCWs who reported being vaccinated against COVID-19 had a 14% higher risk of being seropositive for SARS-CoV-2 IgG compared to those who reported not being vaccinated (aPR (95% CI) = 1.14 (1.03- 1.26)). HCWs who believed to have contracted COVID-19 previously but never tested had an 11% higher risk of being seropositive for SARS-CoV-2 IgG compared to those who thought never to have contracted COVID-19 before (aPR (95% CI) = 1.11 (1.05- 1.16)).

Healthcare workers at ARRH and Mloganzila Hospital had a 9% and 7% higher risk of being seropositive for SARS-CoV-2 IgG respectively compared to HCWs at MNH (aPR (95% CI) = 1.09 (1.05- 1.12)) and (aPR (95% CI) = 1.06 (1.01- 1.13)). The risk of being seropositive for SARS-CoV-2 IgG was 19% higher among laboratory HCWs compared to HCWs at the ED (aPR (95% CI) = 1.21 (1.12- 1.27)). HCWs in Obstetrics and Gynecology had a 19% higher risk of being seropositive for SARS-CoV-2 IgG compared to HCWs at the ED (aPR (95% CI) = 1.19 (1.05- 1.35)). Anesthetists had a 20% higher risk of being seropositive for SARS-CoV-2 IgG compared to HCWs at the ED (aPR (95% CI) = 1.20 (1.10- 1.32)).

Compared to nurses, specialist doctors had a 9% lower risk of being seropositive for SARS-CoV-2 IgG (aPR (95% CI) = 0.91 (0.86 – 0.97)).

HCWs who reported being divorced/ separated had a 13% higher risk of being seropositive compared to those who reported being married/ cohabiting (aPR (95% CI) = 1.13 (1.10- 1.15)) – **Table 3**.

**Table 3:** Factors associated with SARS-CoV-2 IgG seropositivity among HCWs in Dar es Salaam (n=394)

| Variable | cPR (95% CI) | p-value | aPR (95% CI) | p-value |
| --- | --- | --- | --- | --- |
| **Sex** |  |  |  |  |
| Male | Ref |  |  |  |
| Female | 0.97 (0.94- 1.00) | 0.034 | 0.94 (0.90- 0.99) | 0.021 |
| **Age Category** |  |  |  |  |
| 20-35 | Ref |  |  |  |
| 36-50 | 1.03 (0.97- 1.08) | 0.370 |  |  |
| 51-65 | 1.02 (0.98- 1.07) | 0.318 |  |  |
| **Marital Status** |  |  |  |  |
| Married/ Co-habiting | Ref |  |  |  |
| Never Married | 0.94 (0.91- 0.98) | 0.006 | 0.97 (0.96- 0.97) | <0.001 |
| Widowed | 1.01 (0.98- 1.03) | 0.662 | 1.01 (1.00- 1.02) | 0.029 |
| Divorced/ Separated | 1.08 (1.05- 1.12) | <0.001 | 1.13 (1.10- 1.15) | <0.001 |
| **Department** |  |  |  |  |
| ED | Ref |  |  |  |
| Internal Medicine | 1.12 (0.93- 1.36) | 0.233 | 1.15 (0.99- 1.34) | 0.076 |
| OPD | 1.12 (0.96- 1.30) | 0.154 | 1.11 (1.01- 1.23) | 0.032 |
| Surgery, ENT & Dental | 1.08 (0.98- 1.19) | 0.111 | 1.09 (1.00- 1.18) | 0.046 |
| Laboratory | 1.11 (0.93- 1.33) | 0.253 | 1.19 (1.12- 1.27) | <0.001 |
| Obstetrics & Gynecology | 1.17 (1.02- 1.34) | 0.026 | 1.19 (1.05- 1.35) | 0.005 |
| Pediatrics | 1.12 (1.01- 1.25) | 0.034 | 1.09 (1.00- 1.19) | 0.044 |
| Anesthesia | 1.23 (1.05- 1.45) | 0.010 | 1.20 (1.10- 1.32) | <0.001 |
| Others | 1.15 (0.91- 1.44) | 0.250 | 1.10 (0.91- 1.32) | 0.343 |
| **Cadre** |  |  |  |  |
| Nurse | Ref |  |  |  |
| Medical Officer/Dentist | 0.99 (0.94- 1.05) | 0.843 | 0.99 (0.97- 1.00) | 0.147 |
| Specialist | 1.00 (0.94- 1.07) | 0.987 | 0.91 (0.86 – 0.97) | 0.002 |
| Ward attendant | 0.96 (0.92- 0.99) | 0.012 | 1.01 (1.00- 1.03) | 0.086 |
| Lab personnel | 0.96 (0.81- 1.12) | 0.578 | 0.93 (0.77- 1.13) | 0.450 |
| Others | 0.98 (0.89- 1.08) | 0.722 | 0.94 (0.87- 1.01) | 0.106 |
| **Vaccination status** |  |  |  |  |
| Yes | 1.13 (1.02- 1.25) | 0.017 | 1.14 (1.03- 1.26) | 0.013 |
| No | Ref |  |  |  |
| **Previous COVID-19 Experience** |  |  |  |  |
| Never had Covid-19 | Ref |  |  |  |
| Tested Positive for Covid-19 | 1.15 (1.06- 1.24) | 0.001 | 1.12 (0.99- 1.26) | 0.064 |
| Believed had COVID-19 but never tested | 1.12 (1.05- 1.19) | 0.001 | 1.11 (1.05- 1.16) | <0.001 |
| **Site** |  |  |  |  |
| MNH | Ref |  |  |  |
| ARRH | 1.09 (1.02- 1.16) | 0.016 | 1.09 (1.05- 1.12) | <0.001 |
| Mloganzila Hospital | 1.06 (0.99- 1.14) | 0.085 | 1.07 (1.01- 1.13) | 0.025 |

Adjusted for: Sex, marital status, department, cadre, vaccination status, previous COVID-19 experience and site; ED- Emergency Department; OPD- Outpatient Department; ENT- Ear, Nose, and Throat; MNH- Muhimbili National Hospital; ARRH- Amana Regional Referral Hospital; cPR- Crude Prevalence Ratio; aPR- Adjusted Prevalence Ratio; CI- Confidence Interval; SARS-CoV-2- Severe Acute Respiratory Syndrome Corona Virus 2.

## Discussion

The overall seroprevalence of SARS-CoV-2 antibodies among HCWs was high compared to the global prevalence of HCW infection reported around the time of initial study enrolment [59.2% in September 2021 vs 91.8% among HCWs in our study] (18). The prevalence of SARS-CoV-2 among HCWs in other parts of the world ranged from 11.7% in Canada to 27% in the USA (8,19). In sub-Saharan Africa, SARS-CoV-2 prevalence ranged from 34.7% among HCWs in Mozambique to over 50% in Madagascar (20).

With only about a third of participants vaccinated, the high seroprevalence indicates a high infection rate with SARS-CoV-2 among HCWs in Dar-es-Salaam (17). Nearly 87% of unvaccinated HCWs had SARS-CoV-2 antibodies. These high estimates suggest several contributing factors including suboptimal infection control practise among HCWs in Dar es Salaam (20–24).

This study also revealed site and duration-specific differences in SARS-CoV-2 IgG N and S seroprevalence and IgG S alone. Participants enrolled at MNH (between October and November 2021) had relatively lower IgG N and S rates (31.1%) than participants enrolled from Mloganzila and ARRH (83%) (between January and March 2022). These findings are most likely related to the emergence of the new Omicron wave which coincided with the later recruitment period. The high exposure to SARS-CoV-2 among HCWs from Mloganzila and ARRH could also account for the higher IgG S alone among HCWs from MNH despite reported higher vaccination rates at Mloganzila and ARRH.

The high prevalence of SARS-CoV-2 immunity including among those who were never suspected to have COVID-19 or vaccinated indicates significant underreporting of SARS-CoV-2 cases in Tanzania. This was almost certainly due to the inadequate availability of testing of SARS-CoV-2 as fewer than a fifth of the HCWs in this study ever tested for COVID-19 (13). Underreporting of the SARS-CoV-2 cases could have also been due to a high frequency of asymptomatic infection, although it is notable that over two-thirds of the HCWs believed that they had COVID-19 even though they never tested, suggesting that most were symptomatic. Similar reports have been confirmed in other settings in SSA (25).

These findings highlight the importance of testing for infections like COVID-19 in HCWs and other key populations where occupational exposure is high, to ensure mitigation strategies/prevention control measures are appropriately implemented. Estimates that rely on the “confirmed” cases underestimate the burden by a large margin when testing coverage is so low and has limited reliability in planning and allocation of resources (25,26). Our study findings also highlight the need for the rapid development of affordable and easily accessible diagnostics in pandemics such as these, particularly in LMICs. (18).

HCW vaccination rates were low in this setting (37.3%). The uptake of the SARS-CoV-2 vaccine among HCWs in Tanzania is low compared to that of high-income countries but comparable to that of the community in Tanzania (27–29). HCWs who were vaccinated were more likely to have antibodies than those who were not vaccinated. It is well known that SARS-CoV-2 vaccines have been shown to protect against severe forms of the disease (30). As one of the ways to ensure that HCWs are protected during pandemics, vaccination should be prioritized and reasons hindering their uptake and acceptability should be examined and quickly addressed (29).

Anesthetists, and HCWs working in the laboratory and Obstetrics and Gynecology departments were more likely to have SARS-CoV-2 IgGs than HCWs from the EMD. Similarly, ward attendants were more likely to have SARS-CoV-2 IgGs than nurses. These findings likely reflect the higher risk of transmission among some cadres where contact with patients is longer, and who tend to be asymptomatic and infection control practices are less likely to be adhered to (7,31). Another notable finding was that female HCWs were less likely to have SARS-CoV-2 antibodies than male HCWs. Studies have shown that although the decay of SAR-CoV-2 anti-N is similar between males and females, females have a steeper decay of SAR-CoV-2 anti-S compared to males (32).

To our knowledge, this is the first study to determine the sero-prevalence of SARS-CoV-2 among HCWs from different cadres in Tanzania, both clinical and clerical, and the burden of SARS-CoV-2 exposure and infection in this relatively low vaccinated population. There were some limitations. The study was conducted in tertiary-level hospitals located in Dar es Salaam limiting the generalizability of the results to other facilities and regions where transmission dynamics and exposure levels differ. Also, the tests used are limited in their ability to differentiate an active infection from a previous one as well as prior infection vs. vaccination.

In conclusion, the seroprevalence of anti-SARS-CoV-2 IgG among HCWs in this study was high. With only a third of HCWs vaccinated, these findings suggest widespread infection rather than vaccine-induced immunity. The high seropositivity even among those never diagnosed with COVID-19 indicates substantial underreporting, likely due to limited testing availability and potentially high rates of asymptomatic infection. Variations in seroprevalence were observed across hospital sites, the timing of recruitment, and healthcare worker roles, with certain cadres like anesthetists and ward attendants showing higher exposure rates. These findings highlight the critical need for improved infection control in healthcare settings, enhanced testing capacity and accessibility, better vaccination coverage among HCWs, and targeted protective measures for high-risk healthcare cadres.

## Data Availability

Data are available from the corresponding authors for researchers who meet the criteria for access to confidential data. To access data, a third party is also required to sign the Data Transfer Agreement through the National Health Research and Ethics Committee which is the research regulatory body.

NA

## Acknowledgements

We would like to extend our heartfelt gratitude to the Muhimbili National Hospital, Mloganzila National Hospital, Amana Referral Hospital, Muhimbili University of Health and Allied Sciences, Northwestern University and all those who contributed to the successful completion of this study. Our sincere thanks go to the research assistants whose dedication and hard work were instrumental in the data collection process. We also wish to acknowledge the laboratory technicians who played a crucial role in the collection and analysis of the blood samples. Additionally, we are deeply grateful to the healthcare workers who generously agreed to participate in this study. This study would not have been possible without your support and commitment.

## Funding details

*This study was partly supported by Northwestern University’s Global Health Catalyzer Fund and Northwestern’s Global Health Initiative, which is generously supported by Northwestern Medicine Primary and Specialty Care and the Muhimbili University of Health and Allied Sciences*.

## Declarations

All authors declare no conflicts of interest.

## Notes

### Competing Interest Statement

The authors have declared no competing interest.

### Clinical Protocols

NA

### Funding Statement

Yes

### Author Declarations

Approval of conducting research was obtained from the Muhimbili University of Health and Allies Sciences and the National Institute for Medical Research (NIMR/HQ/R.8a/Vol.IX/3774).

## References

1. Lal A, Erondu NA, Heymann DL, Gitahi G, Yates R. Fragmented health systems in COVID-19: rectifying the misalignment between global health security and universal health coverage. Lancet [Internet]. 2021;397(10268):61–7. Available from: 10.1016/S0140-6736(20)32228-5

2. Mehta S, Machado F, Kwizera A, Papazian L, Moss M, Azoulay É, et al. COVID-19: a heavy toll on health-care workers. Lancet Respir Med [Internet]. 2021 Mar;9(3):226–8. Available from: https://linkinghub.elsevier.com/retrieve/pii/S2213260021000680

3. Moyazzem Hossain M, Abdulla F, Rahman A. Challenges and difficulties faced in low- and middle-income countries during COVID-19. Heal Policy OPEN [Internet]. 2022;3(October):100082. Available from: 10.1016/j.hpopen.2022.100082

4. Kana BD, Arbuthnot P, Botwe BK, Choonara YE, Hassan F, Louzir H, et al. Opportunities and challenges of leveraging COVID-19 vaccine innovation and technologies for developing sustainable vaccine manufacturing capabilities in Africa. Lancet Infect Dis [Internet]. 2023;23(8):e288–300. Available from: 10.1016/S1473-3099(22)00878-7

5. Alakija A. Leveraging lessons from the COVID-19 pandemic to strengthen low-income and middle-income country preparedness for future global health threats. Lancet Infect Dis [Internet]. 2023;23(8):e310–7. Available from: 10.1016/S1473-3099(23)00279-7

6. Wang H, Zeng W, Kabubei KM, Rasanathan J, Kazungu J, Ginindza S, et al. The Economic Burden of SARS-CoV-2 Infection Amongst Health Care Workers in the First Year of the Pandemic in Kenya, Colombia, Eswatini, and South Africa [Internet]. Washington; 2023. Available from: www.worldbank.org

7. Costantino C, Cannizzaro E, Verso MG, Tramuto F, Maida CM, Lacca G, et al. SARS-CoV-2 Infection in Healthcare Professionals and General Population During “First Wave” of COVID-19 Pandemic : A Cross-Sectional Study Conducted in Sicily, Italy. Front Public Heal. 2021;9(May).

8. Venugopal U, Jilani N, Rabah S, Shariff MA, Jawed M, Mendez Batres A, et al. SARS-CoV-2 seroprevalence among health care workers in a New York City hospital: A cross-sectional analysis during the COVID-19 pandemic. Int J Infect Dis [Internet]. 2021 Jan;102(102):63–9. Available from: https://linkinghub.elsevier.com/retrieve/pii/S1201971220322402

9. Bitencourt FV, Lia EN, Pauletto P, Martins CC, Stefani CM, Massignan C, et al. Prevalence of SARS-CoV-2 infection among oral health care workers worldwide: A meta-analysis. Community Dent Oral Epidemiol. 2022;(November):1–11.

10. WHO. The impact of COVID-19 on health and care workers: a closer look at deaths. 2021;(September).

11. Mfinanga SG, Mnyambwa NP, Minja DT, Ntinginya NE, Ngadaya E, Makani J, et al. Tanzania’s position on the COVID-19 pandemic Global health and its discontents. Lancet [Internet]. 2021;397(10284):1542–3. Available from: 10.1016/S0140-6736(21)00678-4

12. Hamisi NM, Dai B, Ibrahim M. Global Health Security amid COVID - 19 : Tanzanian government’s response to the COVID - 19 Pandemic. BMC Public Health [Internet]. 2023;1–10. Available from: 10.1186/s12889-023-14991-7

13. Kangwerema A, Thomas H, Knovicks S, Safari J, Diluxe M, Madadi S, et al. The challenge of dearth of information in Tanzania’s COVID-19 response. J Glob Heal Sci. 2021;3(2):2–5.

14. MOHCDGEC. COVID-19 SITUATION REPORT: NO 18 From 08th to 14th January, 2022 [Internet]. MINISTRY OF HEALTH, COMMUNITY DEVELOPMENT, GENDER, ELDERLY AND CHILDREN. 2022. p. 6. Available from: https://www.moh.go.tz/report

15. Tarimo CS, Wu J. The first confirmed case of COVID-19 in Tanzania : recommendations based on lesson learned from China. Trop Med Health. 2020;48(25):0–2.

16. Mfinanga SG, Gatei W, Tinuga F, Mwengee WMP, Yoti Z, Kapologwe N, et al. Tanzania’s COVID-19 vaccination strategy: lessons, learning, and execution. Lancet [Internet]. 2023;401(10389):1649. Available from: 10.1016/S0140-6736(23)00723-7

17. Castilla J, Lecea Ó, Salas CM, Quílez D, Miqueleiz A, Trobajo-sanmartín C. Seroprevalence of antibodies against SARS-CoV-2 and risk of COVID-19 in Navarre, Spain, May to July 2022. Eurosurveillance [Internet]. 2022;27(33):0–6. Available from: 10.2807/1560-7917.ES.2022.27.33.2200619

18. Bergeri I, Whelan MG, Ware H, Subissi L, Nardone A, Lewis HC, et al. Global SARS-CoV-2 seroprevalence from January 2020 to April 2022: A systematic review and meta-analysis of standardized population-based studies. PLoS Med [Internet]. 2022;19(11):1–24. Available from: 10.1371/journal.pmed.1004107

19. Brousseau N, Morin L, Ouakki M, Savard P, Quach C, Longtin Y, et al. SARS-CoV-2 seroprevalence in health care workers from 10 hospitals in Quebec, Canada: a cross-sectional study. Cmaj. 2021;193(49):E1868–77.

20. Abrokwa SK, Ravaoarisoa L, Briesemeister V, Andrianasolo RL, Andrianarivelo AM, Müller SA, et al. A multi-site cross-sectional study on the burden of SARS-CoV-2 in healthcare workers in Madagascar. PLoS One. 2024;19(10 October):1–15.

21. Laverdure S, Kazadi D, Kone K, Callier V, Dabitao D, Dennis D, et al. SARS-CoV-2 seroprevalence in vaccine-naïve participants from the Democratic Republic of Congo, Guinea, Liberia, and Mali. Int J Infect Dis. 2024;142.

22. Madinga J, Mbala-Kingebeni P, Nkuba-Ndaye A, Baketana-Kinzonzi L, Matungulu-Biyala E, Mutombo-Lupola P, et al. COVID-19 seroprevalence cohort survey among health care workers and their household members in Kinshasa, DR Congo, 2020–2022. J Heal Popul Nutr. 2024;43(1):1–13.

23. Mostafa A, Kandil S, El-Sayed MH, Girgis S, Hafez H, Yosef M, et al. SARS-CoV-2 seroconversion among 4040 Egyptian healthcare workers in 12 resource-limited healthcare facilities: A prospective cohort study. Int J Infect Dis. 2021;104:534–42.

24. Nyaulingo BC, Mhimbira FA. Facilitators and barriers in implementation of active TB drug safety monitoring and management (aDSM) in programmatic management of drug resistance TB in Dar es Salaam region. PLoS One. 2023;18(9):e0291225.

25. Ingelbeen B, Cumbane V, Mandlate F, Barbé B, Nhachungue SM, Cavele N, et al. Mild and moderate COVID-19 during Alpha, Delta and Omicron pandemic waves in urban Maputo, Mozambique, December 2020-March 2022: A population-based surveillance study. PLOS Glob Public Heal. 2024;4(8):1–14.

26. Kluytmans-van den Bergh MFQ, Buiting AGM, Pas SD, Bentvelsen RG, van den Bijllaardt W, van Oudheusden AJG, et al. Prevalence and Clinical Presentation of Health Care Workers With Symptoms of Coronavirus Disease 2019 in 2 Dutch Hospitals During an Early Phase of the Pandemic. JAMA Netw Open [Internet]. 2020 May 21;3(5):e209673. Available from: https://jamanetwork.com/journals/jamanetworkopen/fullarticle/2766228

27. Madran B, Kayı İ, Beşer A, Ergönül Ö. Uptake of COVID-19 vaccines among healthcare workers and the effect of nudging interventions: A mixed methods study. Vaccine [Internet]. 2023 Jul;41(31):4586–93. Available from: https://linkinghub.elsevier.com/retrieve/pii/S0264410X23006886

28. Noushad M, Rastam S, Nassani MZ, Al-Saqqaf IS, Hussain M, Yaroko AA, et al. A Global Survey of COVID-19 Vaccine Acceptance Among Healthcare Workers. Front Public Heal. 2022;9(February):1–12.

29. Msuya SE, Manongi RN, Jonas N, Mtei M, Amour C, Mgongo MB, et al. COVID-19 Vaccine Uptake and Associated Factors in Sub-Saharan Africa: Evidence from a Community-Based Survey in Tanzania. Vaccines. 2023;11(2):1–13.

30. Wang RC, Murphy CE, Kornblith AE, Hohenstein NA, Carter CM, Wong AHK, et al. SARS COV-2 anti-nucleocapsid and anti-spike antibodies in an emergency department healthcare worker cohort: September 2020 – April 2021. Am J Emerg Med [Internet]. 2022;54(December 2020):81–6. Available from: 10.1016/j.ajem.2022.01.055

31. Hossain A, Nasrullah SM, Tasnim Z, Hasan MK, Hasan MM. Seroprevalence of SARS-CoV-2 IgG antibodies among health care workers prior to vaccine administration in Europe, the USA and East Asia: A systematic review and meta-analysis. EClinicalMedicine. 2021;33.

32. Grzelak L, Velay A, Madec Y, Gallais F, Staropoli I, Schmidt-Mutter C, et al. Sex Differences in the Evolution of Neutralizing Antibodies to Severe Acute Respiratory Syndrome Coronavirus 2. J Infect Dis. 2021;224(6):983–8.

